# Beyond Accuracy in Small Open-Source Medical Large Language Models for Pediatric Endocrinology

**DOI:** 10.1101/2025.08.26.25334485

**Authors:** Vanessa D’Amario, Randy Daniel, Dhruv Edamadaka, Nitya Alaparthy, Joshua Tarkoff

**Affiliations:** Department of Decision Sciences, H. Wayne Huizenga College of Business and Entrepreneurship, Nova Southeastern University; Fort Lauderdale (USA); Department of Health Informatics, Dr. Kiran C. Patel College of Osteopathic Medicine, Nova Southeastern University; Fort Lauderdale (USA); Nicklaus Children’s Hospital; Miami (USA)

**Author notes:** **Corresponding author:** Vanessa D’Amario.

**Keywords:** Large Language Models, Open-Source, Stochastic Inference, Prompting, Robustness, Pediatric Endocrinology

## Abstract

**Objective:** Small open-source medical large language models (LLMs) offer promising opportunities for low-resource deployment and broader accessibility. However, their evaluation is often limited to accuracy on medical multiple choice question benchmarks, and lacks evaluation of consistency, robustness, or reasoning behavior.

**Materials and Methods:** We evaluate six small open-source medical LLMs (HuatuoGPT-o1 ^1^, Diabetica-7B ^2^, Diabetica-o1 ^2^, Meditron3-8B ^3^, MedFound-7B ^4^, and ClinicaGPT-base-zh ^5^) in deterministic settings, where we examine how prompt variation and removal of option labels affect models output. In stochastic settings, we evaluate the variability of models responses and investigate the relationship between consistency and correctness. Lastly, we evaluate self-assessment bias by testing whether high-performing models can recognize the correct reasoning path when presented with gold-standard explanations. The response evaluation consists of a mix of human reviews and pediatric endocrinology expert.

**Results:** HuatuoGPT-o1-8B achieved the highest score with 32 correct responses of the 91 cases considered. All models exhibited high sensitivity to prompt phrasing (maximum level of agreement Cohen’s *κ* = 0.55) and to label removal (highest Cohen’s *κ* = 0.35). The results show that high consistency across the model response is not an indicator of the correctness of the model, although HuatuoGPT-o1-8B showed the highest consistency rate. When tasked with selecting correct reasoning, both HuatuoGPT-o1-8B and Diabetica-o1 exhibit self-assessment bias and dependency on the order of the candidate explanations. Expert review of incorrect reasoning rationales identified a mix of clinically acceptable responses and clinical oversight.

**Discussion:** Although none of the evaluated models fall short of demonstrating deep expertise in pediatric endocrinology, HuatuoGPT-o1-8B demonstrated the highest robustness to input variability and highest stability across variation of the hyperparameters used for inference.

**Conclusion:** This work underscores the limitations of relying solely on accuracy for evaluating medical LLMs and proposed a broader diagnostic framework to understand potential pitfalls in real-world clinical decision support.

## 1 INTRODUCTION

Large Language Models (LLMs) hold significant potential in healthcare, particularly to improve chronic disease management and clinical decision support ^6,7^. High-performance medical LLMs have showed strong results on different aspects of clinical care, from diagnostics to patient interfacing ^4,8,9^. However, these models are often large, exceeding 70 billions parameters, and proprietary, making them inaccessible for independent validation or adaptation. They become impracticable solutions in resource-constrained or low-income healthcare settings.

Differently, small open-source medical LLMs offer the possibility of lightweight, locally deployable tools that could reduce barriers to access. A critical question to address is understanding if these models, as released and without fine-tuning, encode sufficient clinical knowledge to support real-world use.

Pediatric endocrinology presents a unique testbed for the evaluation of medical LLM. Pediatric endocrinology is facing a growing patients volume, longer waiting time, and larger range of conditions in the last years ^10,11^. At the same time, access to pediatric endocrinology expertise is limited in many regions ^11^, creating a potential role for AI-driven decision support.

In this study, we explore this question by evaluating a selection of small (under 10 billion parameters) previously fine-tuned open-source medical LLMs in pediatric endocrinology.

Given the lack of evaluation of medical LLMs in this field, our work fills this gap by systematically assessing model behavior on a curated set of pediatric endocrinology cases, offering insights into both the capabilities and limitations of these tools in a real-world clinical subspecialty.

To evaluate model performance in pediatric endocrinology, we leverage Multiple Choice Questions (MCQs) drawn from real-world clinical content. MCQs are a widely used strategy for evaluating medical LLMs, particularly small, domain-specific models. While convenient and well-structured for automated scoring and scalable benchmarking, MCQ-based evaluation has well-documented limitations. Question difficulty varies widely, distractors may lack clinical plausibility, and accuracy alone often obscures reasoning quality, robustness, or consistency. Recent studies outside of the healthcare context ^12,13^ have also highlighted response instability among LLMs in MCQ settings and self-assessment bias ^14–16^.

Despite these challenges, MCQs remain one of the most accessible formats for systematic model evaluation. In this work, we embrace their practicality while addressing their shortcomings: we propose a multidimensional evaluation framework that goes beyond standard accuracy-based evaluation. We evaluate six small and assess *models consistency*, by comparing decisions across prompt variations, including syntactical variations and the removal of answer option labels. We also evaluate *response consistency and correctness* across stochastic sampling, where the degree of stochasticity of the inference process is determined by different temperature values. Lastly, we evaluate the presence of *self-assessment bias*, by presenting models with a choice between their own rationale and a gold-standard clinical explanation.

This multidimensional analysis provides a structured diagnostic of small medical LLM readiness and exposes failure modes that accuracy metrics alone would not be able to capture. Our goal is not only to compare models but to define critical evaluation criteria for trustworthy, accessible LLMs in medicine. In doing so, we highlight which models show the greatest promise for practical clinical integration and identify areas requiring further improvement.

## 2 MATERIALS AND METHODS

### 2.1 Multiple Choice Questions from Pediatric ESAP 2021-2022

To evaluate the reasoning capabilities of small opensource medical LLMs we rely on the Pediatric Endocrine Self-Assessment Program 2021-2022 (ESAP) ^17^. The program is a question bank containing 100 among clinical cases and factual questions based on clinical guidelines. Clinical cases-items present a patient scenario, including relevant medical history, examination findings, and laboratory results, and typically ask to determine the most likely diagnosis or the best course of action in patient management. Other more factual items focus on guideline-based knowledge such as indications for specific medications, appropriate monitoring strategies, or recommended referral to a subspecialist. Each ESAP item includes a description, a question, and five possible answers, educational objective, correct answer, and clinical explanation for each case. Drawing on the expertise of leading pediatric endocrinologists, the program is highly technical and aimed at medical specialists. The program’s learning objectives include recognizing clinical features of endocrine, growth, and metabolic disorders; applying current diagnostic and treatment options; identifying risk factors; and evaluating endocrine manifestations of systemic diseases. As such, we considered it as an optimal starting point for knowledge assessment in pediatric endocrinology.

We excluded nine ESAP items from the analysis, since those rely on images and graphs. For each clinical case described, we extracted the following fields: `<description>, <question>, <options>, <correctanswer>, and <explanation>`.

### 2.2 Small Open-Source Medical LLMs

We evaluate six small-sized, open-source medical LLMs designed for biomedical reasoning.

*HuatuoGPT-o1* ^1^ is based on the LLaMA 3.1 8B architecture and is trained using continued pre-training and supervised fine-tuning in a bilingual corpus of 1.1TB (Chinese and English), with benchmarks including MedQA, MedMCQA, CMB, CMExam, and CMMLU.

*Diabetica-7B* ^2^ uses the Qwen2-7B-Instruct model, fine-tuned through LoRA and self-distillation on public medical QA sets, SFT datasets, and internal diabetes-specific data, evaluated through MCQs, fill-in-the-blank and dialogue tasks. *Diabetica-o1* is the result of self-distillation of Diabetica-7B.

*Meditron3-8B* ^3^, using LLaMA 3.1-8B, underwent a continuous pre-training and supervised instruction fine-tuning in PubMed Central, validated medical textbooks, and more than 46,000 clinical practice guidelines, and was tested on MedQA, MedMCQA, and PubMedQA.

*MedFound-7B* ^4^ and *ClinicalGPT-based-zh* ^5^ are both based on the BLOOM-7B. The latter was developed through supervised fine-tuning and reinforcement learning using a range of Chinese electronic health records, multi-turn patient-doctor dialogues, standardized medical examination questions, and question-answer pairs derived from structured medical knowledge graphs.

### 2.3 Experimental Design

To assess the stability and reasoning capabilities of small open-source medical LLMs, we prepared our dataset from the Pediatric ESAP 2021-2022 as a stepping-stone in the design of three main experiments. We describe the inferential setup for all the experiments in App. B.1.

#### 2.3.1 Experiment 1: LLMs Accuracy and Response Stability to Prompt Variation

We evaluate the performance of the models under deterministic conditions (temperature *T* = 0). Given the ESAP item, including description question and options, each model is instructed to select the correct option and provide supporting clinical reasoning. Three prompt strategies are tested: (1) prompt A (Appendix A.1), (2) prompt B, which slight syntactical variations to prompt A and is used to assess prompt sensitivity; (3) prompt A without letter token, which is identical to prompt A, but excludes the option letter (A,B,C,D,E) from the <options> field, separating each option with a semicolon.

The main goal of this experiment is to measure model accuracy on forced-choice selection and evaluate medical LLMs stability to prompt variations, using *κ* Cohen’s coefficient.

#### 2.3.2 Experiment 2: Stability in non-deterministic setting

We examine the reliability of the prediction under stochastic decoding, for increasing temperature values. Despite ^18,19^ found stable performance of commercial models (ChatGPT, Llama 70B) across different temperatures, recent studies have showed how model performance degrades for LLMs of limited size (*<* 13 B) over *T* ≥1, especially for tasks that require instruction following ^20^. In our experiments, we evaluate the model in three different settings *T* = [0.3, 0.6, 1.0]. We focus on the models that perform the best in Experiment 1. We define *consistency* as the frequency of *majority vote* and *correctness* as the frequency of correct answers in ten runs. This setup allows us to assess whether high-confidence predictions consistently correspond to correct answers. We also determine the model stability at different stochastic regimes and the best performing model at different regimes.

#### 2.3.3 Experiment 3: Self-Bias and Discerning Gold-Standard Reasoning

In this setup, we provide the LLM with the ESAP item description, question, and multiple choice options. We ask the LLM to select the most clinically correct solution between two candidate explanations: (1) the model’s own output from Experiment 1, and (2) a gold-standard explanation authored by ESAP instructors (prompt in App. A.2). We additionally ask the LLM to output a degree of agreement between the two explanations on a scale from 0 to 5. To evaluate the presence of positional bias, we prompted the model in two ways: once with the gold-standard explanation listed first and the model’s reasoning second, and then with reversed order. We focus on the ESAP items where the model selected an incorrect response in Experiment 1, since those necessarily diverge from the indicated best answer. Incorrect might reflect factually wrong statements, as well as overly cautious management plan, which do not align with clinical expectations.

Given the time-consuming nature of this task, a pediatric endocrinology is asked to review the clinical reasoning of the best-performing model on items that were consistently answer incorrectly, independent of the position bias. The evalution is based on one of the items of the CLEVER framework proposed for clinical reasoning, in ^4^, using a Likert scale from 1 to 5. Clinical reasoning is defined as the alignment of the LLM’s content with the diagnostic reasoning process used in medical practice.

#### 2.3.4 Output Evaluation

In our initial design, we planned to automate the output evaluation using GPT-4o, but we abandoned this approach due to lack of consistency in GPT-4o responses. The output evaluation is conducted separately by three non-medical reviewers. Beyond the five standard ESAP options (A-E), we include two further categories: *Multiple Selection* and *Hallucinate or None*. We resume to *Multiple Selection* whenever a model selects two or more options. In cases where the model response includes conflicting or ambiguous selections (*e*.*g*., indicating one answer, then contradicting itself), the output is marked as *Hallucinate or None. Hallucinate or None* reflects the difficulty of non-endocrinology reviewers to draw a line between valid but non-assertive medical verbiage from subtle medical errors.

For instances where non-medical reviewers could not attribute one of the categories, two clinical experts, one in emergency medicine and one in pediatric endocrinology, are consulted to assist with classification.

The code to reproduce LLMs responses and results is available at https://github.com/vanessadamario/SmallMedLLMs-PedEndo.

## 3 RESULTS

Before showing the results for each of the experiments, we comment on the overall performance and hallucinations observed during output evaluation.

Among the models, *HuatuoGPT-o1-8B, Diabetica-7B*, and *Meditron3-8B* exhibited the most consistent reasoning patterns. Their responses generally adhered to the multiple-choice format, often discussing the given options and weighing their clinical likelihood before reaching a conclusion. HuatuoGPT-o1-8B occasionally reversed this order, presenting a final answer first, followed by a discussion of each option, as asked in the prompt. In contrast, *Diabetica-o1* produced outputs that were more difficult to interpret. While the initial portion of the response is often coherent, the output frequently degenerates into simulated dialogues between a fictitious user and an assistant. These interactions included follow-up questions and, in some cases, newly generated MCQs. Hallucinations also included emojis, Chinese ideograms, LaTeX or Python code fragments, or statistical references unrelated to the task. For the evaluation, we considered the first part of the output, and discarded what was unrelated to the analysis of the clinical scenario.

The output from *MedFound-7B* also showed signs of degeneration. The model frequently hallucinated content, and when not hallucinating, it was hesitant to provide a definitive answer. Instead, it tended to suggest a wide array of additional diagnostic tests before committing to a conclusion. The *ClinicalGPT-base-zh* model produced shorter responses, typically only a few sentences, and rarely addressed all options. It often displayed a strong preference for a single option without discussing alternatives. For quantitative evaluations of option distributions, see App. C.

### 3.1 LLMs Performance, Prompt Stability, and Agreement

In Table 1, we report the performance of the six LLMs on the ESAP Program items. The left panel lists each model’s number of correct responses and its usability rate, calculated as the percentage of outputs yielding a valid single-option selection. The middle panel shows performance under prompt B. The agreement column reports (1) Cohen’s *κ* coefficient of agreement, while the numbers in parentheses show the number of agreed cases between prompts A and B (2), and the number of agreed and correctly answered cases (3). The right panel mirrors these metrics for prompt A when option letters are omitted.

**Table 1.** *Legend*. **#correct**: number of correct responses out of 91 ESAP items; **usab**: percentage of usable responses, where the decision falls in one of the five ESAP options; **agreement** indicated as <C>(<D>|<E>): <C>=Cohen’s *κ*, <D>= number of decisions where there is agreement to prompt A, <E>=both agreement and correct. Each row shows the performance For a different model. Performance and agreement are reported for a prompt (prompt B) similar to prompt A (center side of the table) and for a prompt identical to prompt A without letter token (right side).

| **Model** | Prompt A | Prompt B (*similar to prompt A*) |  | Prompt A Without Letter Token |  |
|---|---|---|---|---|---|
|  | #correct(usab) | #correct(usab) | agreement | #correct(usab) | agreement |
| **HuatuoGPT-o1-8B**<sup>1</sup> | **32**(**100**) | **32**(**100**) | **0.55**(**58**\|**23**) | **30**(**96.7**) | **0.35**(**44**\|**19**) |
| Diabetica-o1<sup>2</sup> | 30(100) | 31(100) | 0.40(47\|17) | 25(94.5) | 0.35(44\|16) |
| Diabetica-7B<sup>2</sup> | 27(98.9) | 29(100) | 0.63(64\|23) | 25(87.9) | 0.38(46\|16) |
| Meditron3-8B<sup>3</sup> | 30(97.8) | −(−) | − | 31(90.1) | 0.19(32\|15) |
| MedFound-7B<sup>4</sup> | 4(18.6) | 10(34.1) | −0.18(5\|1) | 4(22.0) | −0.20(4\|2) |
| ClinicalGPT-base-zh<sup>5</sup> | 18(79.1) | 18(94.5) | 0.15(29\|6) | 18(79.1) | 0.18(31\|10) |

None of the models approached 91 correct responses, indicating limited accuracy. *HuatuoGPT-o1-8B* emerges as the model with highest number of correct responses across prompt A and prompt B. Although HuatuoGPT-o1-8B is slightly more accurate than Diabetica-o1, a closer look at the LLMs outputs reveals important differences regarding hallucinations and additional content appearing in output for Diabetica-o1. A breakdown on Diabetica-o1 performance shows that under prompt A and B the model outputs respectively only 12 and 13 fully interpretable answers, the output contains emojis in 12 and 14 cases, the output is truncated in 34 and 45 cases, the output contains LaTeX, Python, or Markdown code in 27 and 23 cases, and Chinese characters in 3 cases, across the two prompts. As expected, Diabetica-7B performs slightly worse than Diabetica-o1. Meditron3-8B shows relatively high performance compared to other models for prompt A, but it fails to produce an output under prompt B. Models with lower overall performance, such as ClinicalGPT-base-zh, exhibit close to chance performance (chance = 18.2 correct answer) and a strong bias toward a particular response (often option A). For further discussion, refer to App. C. The level of agreement of the model’s answer between the prompt A and prompt B is generally low, showing a high dependence from syntactical chages. Only Diabetica-7B and HuatuoGPT-o1-8B exceed 0.4 Cohen’s *κ* coefficient, showing a moderate to good level of agreement.

The results evaluating LLM performance under removal of option letters highlight an even stronger sensitivity bias toward specific options. In this setting, all models exhibit lower Cohen’s *κ* coefficients compared to the syntactic variation condition.

### 3.2 Stability in Non-Deterministic Setting

HuatuoGPT-o1-8B, Diabetica-o1, Diabetica-7B, and Meditron3-8B emerged as best performing models from Experiment 1 and were used for follow-up analysis assessing the stability of their responses in the nondeterministic setting. Each LLM was presented with prompt A on ten independent runs.

In Figure 1a, we excluded ESAP items for which the model fails to produce a majority class and report how often a model selected the correct answer versus how often the same model selected its majority class. An ideal model would show all the density concentrated in a single blob at the top-right corner of the plot. In practice, we observe that the tested models deviate from this ideal behavior following different trends. HuatuoGPT-o1-8B shows the highest overall number of consistently correct responses across ten runs for all temperatures. HuatuoGPT-o1-8B is the only model for which we observe an increasing rate of correct responses over incorrect. By contrast, Diabetica-o1 and Diabetica-7B exhibit a sharp decline in model consistency for increasing temperature, with Diabetica-7B showing strong consistency for incorrect responses at *T* = 0.3. Meditron3-8B becomes more reluctant in providing a final answer at higher temperature values. At *T* = 1.0, Meditron3-8B is the only case where the majority vote corresponds to the *Hallucination /None*, for 71 out of 91 outputs, explaining the high density towards the left side of the plot. We report frequencies of the majority class in Supplemental Materials (Tables 6, 7, 8).

**Figure 1.**
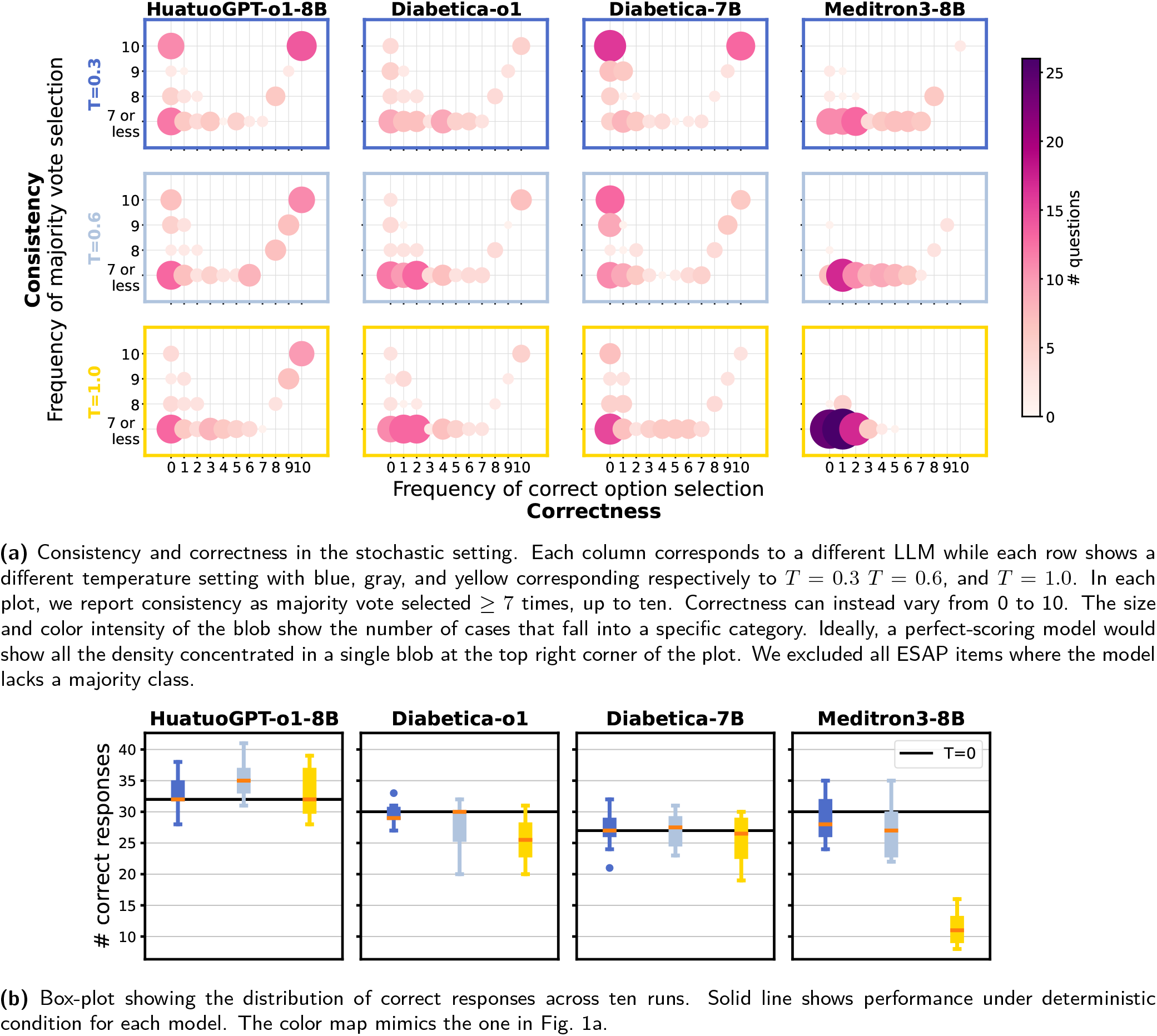

In Table 2, we further examine cases with consistency equal to 10 to assess the effect of temperature on model correctness. For all models except Diabeticao1, overall consistency decreases as temperature increases. HuatuoGPT-o1-8B shows the most stable behavior across different temperatures, where interestingly, the temperature increase affects more abruptly the consistency of wrong answers, leaving a relatively higher number of correct answers still highly consistent.

**Table 2.** Number of ESAP items where the model consistently selected the same option 10 out of 10 runs. The responses are divided in correct and incorrect. C (white background) and I (gray background) indicate respectively count for correct and incorrect responses with maximum consistency.

|  | *T* = 0.3 |  | *T* = 0.6 |  | *T* = 1.0 |  |
|---|---|---|---|---|---|---|
|  | correct | incorrect | C | I | C | I |
| HuatuoGPT-o1-8B | 14 | 11 | 11 | 7 | 10 | 4 |
| Diabetica-o1 | 5 | 4 | 7 | 3 | 5 | 3 |
| Diabetica-7B | 13 | 16 | 6 | 13 | 3 | 7 |
| Meditron3-8B | 2 | 0 | 0 | 0 | 0 | 0 |

These results highlight that consistency, as reflected by output frequency across runs, does not necessarily map to correctness leading to potential risks of over-relying on output frequency as a proxy for reliability. More detailed results for single case consistency and correctness across at different temperatures are reported in the supplementary material (see Fig. 3) for the four models tested.

### 3.3 Discerning Gold-Standard Reasoning

We retrieved HuatuoGPT-o1-8B and Diabetica-o1 outputs from Experiment 1 (prompt A and prompt B).

Our analysis focuses on the cases where the model responded incorrectly: 59 for both prompts in HuatuoGPT-o1-8B, and 61 and 60 for Diabetica-o1 under prompts A and B, respectively (see Table 1. Figure 2

**Figure 2.**
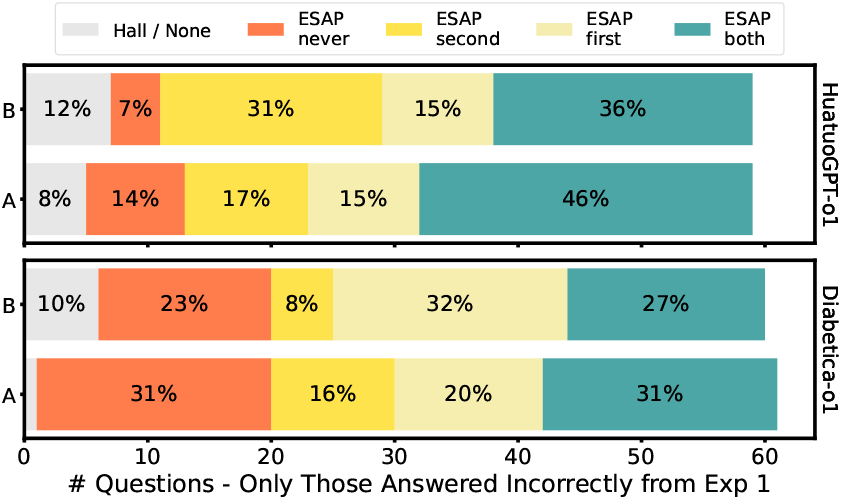
HuatuoGPT-o1-8B and Diabetica-o1 selection of gold-standard explanation against their own reasoning under prompt A and prompt B. Each bar reports the percentage of times each model selected the gold standard explanation, regardless of the presented order (*ESAP both* in green), if only in one of the two positions (*ESAP first* in khaki or *ESAP second* in yellow), in neither (*ESAP never* in red). Cases where a model did not make a valid selection, either by failing to choose between the two explanations or by introducing a new and unrelated option are indicated as *Hallucinate /None*, in gray.

HuatuoGPT-o1-8B shows higher preference for the gold standard explanation than Diabetica-o1. Under prompt A (and B), the HuatuoGPT-o1-8B selected the gold standard explanation in both positions for 27 (21) cases, selected it in only one position for 19 (26) cases, and it failed to select it in either position for 8 (4) cases. There were 5 (7) cases that could not be classified due to the ambiguous or irrelevant output. In comparison, Diabetica-o1 selected the gold standard explanation in both positions for 19 (16) cases, in only one position for 22 (24) cases, and never for 19 (14) cases, with 1 (6) non-classifiable responses under prompt 1 (and 2). These results suggest that HuatuoGPT-o1-8B exhibits a stronger ability to recognize clinically sound reasoning, and in some cases, reformulate its own reasoning to align with the correct explanation.

A pediatric endocrinology expert reviewed the subset of HuatuoGPT-o1-8B outputs where the model selected for the *ESAP never* cases. The physician reported for a total of 12 evaluated ESAP items (see App E.2). For more detailed clinical comments and comparison with HuatuoGPT-o1-8B output, see Table 3. In general, the expert noted how HuatuoGPT-o1-8B often overlooked at key biochemical findings in the ESAP item description, details that would typically lead a clinician to immediately consider or rule out specific options. However, only two of the twelve reviewed cases contain factually incorrect statements.

**Table 3.** Endocrinologist evaluation of HuatuoGPT-o1-8B output on questions where model failed to identify the correct reasoning. Responses are evaluated based on clinical reasoning on a Likert scale from 1 to 5.

| **Prompt** | **Case** | **Score** | **Main Observations** |
|---|---|---|---|
| 1 | 27 | 3 | Missed Factual Details and Key Diagnosis |
|  | 36 | 4 | Acceptable Reasoning |
|  | 37 | 4 | Acceptable Reasoning |
|  | 57 | 4 | Acceptable Reasoning and Clinical Deviation |
|  | 60 | 3 | Partially Correct |
|  | 71 | 2 | Misclassification of Risk |
|  | 75 | 2 | Missed Factual Details |
|  | 94 | 3 | Incomplete Reasoning |
| 2 | 27 | 2 | Wrong Statements<br>Missed Factual Details |
|  | 34 | 2 | Wrong Statements |
|  | 49 | 2 | Misinterpreted Labs |
|  | 94 | 2 | Factual Recall Failure |

Additional considerations on the agreement between the two explanation, as well as differences between the evaluation provided by the pediatric endocrinologist and HuatuoGPT-o1-8B are presented in App. E.1 and App. E.2. These findings suggest that higher agreement levels in HuatuoGPT-o1-8B’s responses are consistent to the model’s tendency to overlook key clinical information for certain items. Interestingly, the scores distributions, as provided by HuatuoGPT-o1-8B, appear more spread when the model recognizes the ESAP explanation as best at either positions, compared to the case where the model never selected the ESAP explanation (see Fig. 4). This shows coherence between the behavior (selection of ESAP choice at both position) and evaluation of quality response. We do not observe a similar behavior for Diabetica-o1 (see Fig. 5).

**Figure 3.**
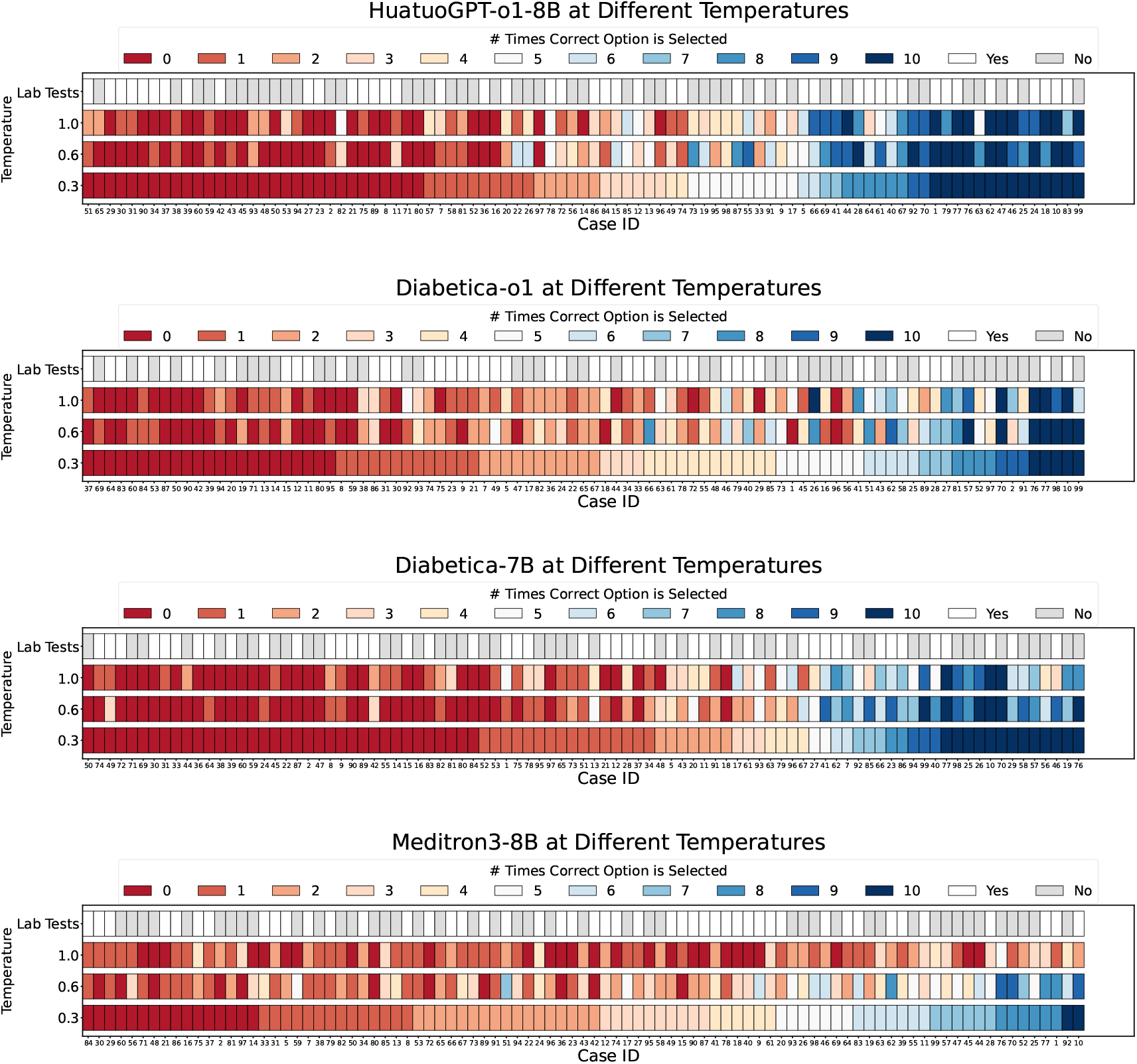
Graphical representation of models consistency and correctness at different temperature values.

**Figure 4.**
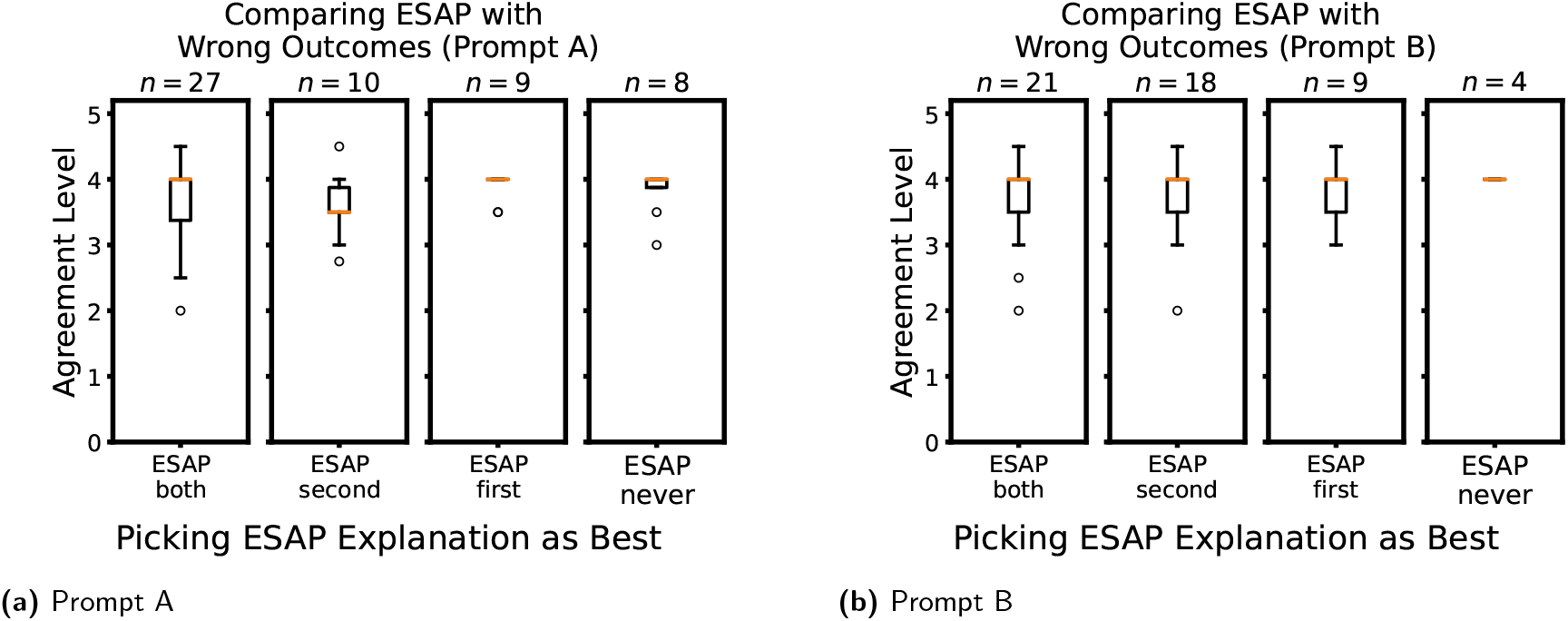
Distribution of agreement scores for **HuatuoGPT-o1-8B**.

**Figure 5.**
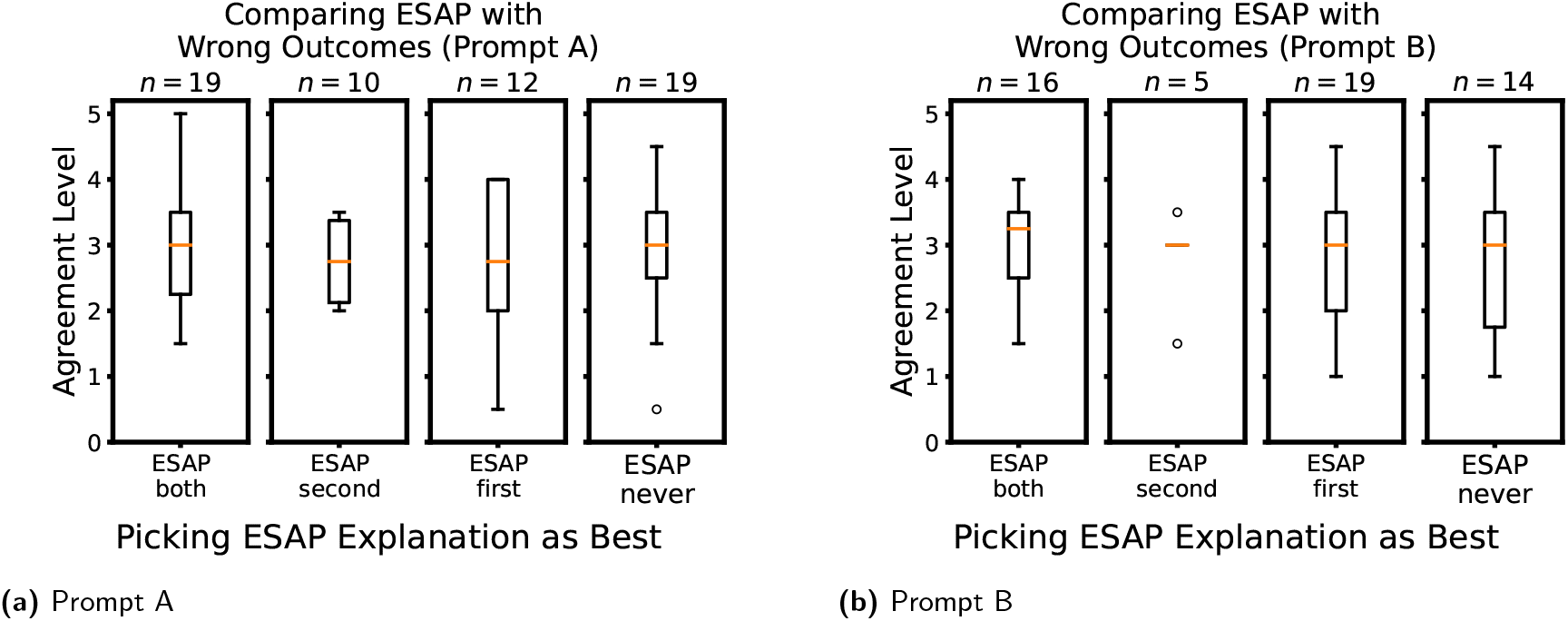
Distribution of agreement scores for **Diabetica-o1**.

## 4 DISCUSSION

### 4.1 Interpretation of Findings

While the tested small open-source medical LLMs exhibit similar overall trends when looking solely at the number of correct answers (Experiment 1, prompt A), the evaluated models largely differ in the quality and consistency of their output and their behavioral patterns under stress-testing. We argue that model evaluation should not rely solely on aggregate performance metrics, but should include finer-grained behavioral diagnostics. These models, despite fine-tuned on medical corpora and MCQs, show a strong bias toward option letters, which becomes particularly problematic in the tested context, suggesting that superficial cues can override semantic reasoning.

As expected, increasing temperature values led to a degradation in model performance, reflecting higher creativity and reduced output stability under stochastic sampling ^18,20^. Although some variation is inherent in LLM ^21^, the degree of inconsistency varies greatly between models, suggesting that HuatuoGPT-o1-8B could be a better fit to clinical tasks than its counterpart. Nonetheless, the results suggest that over-relying on response frequency as a measure of correctness would be misleading. In clinical contexts, such behavior could translate into models persistently recommending inappropriate actions, with the risk of increasing potential harm.

Lastly, even the LLM with strongest overall performance, HuatuoGPT-o1-8B, shows a tendency to favor its own explanations over a provided gold-standard rationale, even when its reasoning is incorrect. The pediatric endocrinologist evaluation in Experiment 3 further revealed that in some cases LLMs output could be acceptable though not strictly correct, whereas in other situations the model made critical errors.

### 4.2 Limitations

First, this work focused on prompt variations, which is proved to affect LLM responses even for large models ^22–24^ and temperature changes ^18,20^, as these parameters have more widely been discussed and directly linked to reasoning consistency and stochastic robustness However, other inference hyperparameters were held constant throughout all the experiments, as detailed in the appendix, and those parameters could possibly have an effect on the model performance. While a more extensive hyperparameter exploration would be valuable, such undertaking remains extremely time-consuming, requiring approximately 60 minutes for the evaluation 91 cases by non-experts.

Additionally, while MCQs provide a scalable framework for benchmarking, our results highlight that LLMs are highly sensitive to prompting. Supplementary experiments compare the output of two technically equivalent inference pipelines (see App. F). Those results show that even routines intended to perform the same task can yield different final option selection. This qualitative evaluation highlights deeper limitations of MCQ-based evaluation methods for assessing LLMs.

Lastly, we acknowledge that non-expert reviewers may overlook details of medical relevance or subtle hallucinations. Nonetheless, our hybrid strategy mixing non-expert reviews with targeted expert evaluation on responses of difficult interpretation offers a practical compromise between rigor and feasibility. A deeper qualitative assessment of the medical content across all model outputs would be extremely valuable but remains prohibitively resource expensive.

### 4.3 Implications and Future Work

None of the tested small open-source models demonstrated sufficient reliability for clinical decision-making “as-is.” Our findings underscore the need for external knowledge integration, fine-tuning, or retrieval-augmented architectures to support more trustworthy outputs, which we aim to pursue in the next phases of our work in order to support the use of these methods in low-resource settings.

Our experiments emphasize that minor changes in question formatting, prompt pipelines, or output constraints can yield divergent responses, even when semantically equivalent. These results raise concerns about the reliance on MCQ formats for LLM evaluation and suggest the importance of alternative approaches such as scenario-based assessment or free-text justification grading. OpenAI recent release of HealthBench ^25^ represents a potential area of exploration.

## 5 CONCLUSION

Our evaluation of small open-source medical LLMs highlights the need to look beyond traditional accuracy metrics and consider other factors such as prompt sensitivity, response stability, and reasoning quality. These dimensions remain largely overlooked in current benchmarking practices, in part due to the deep-rooted challenge of automating reasoning evaluation in highly specialized fields such as pediatric endocrinology. Despite low absolute performance, difficult to contextualize without a human baseline, our analysis reveals how variability in model outputs underscore the need for rigorous and interpretability-focused evaluation frameworks that go beyond raw accuracy.

## Data Availability

Data will be available after publication. Code is available for reproducibility.

## 6 ACKNOWLEDGEMENTS

This work was supported by the National Artificial Intelligence Research Resource Pilot (NAIRR Grant No. 240463) and the NSU Presidential Research Grant (PRG-NSU 2026). We gratefully acknowledge Paul Gerbino and Daniel Schaible of the Endocrine Society for their interest in this study, and Veronica Tozzo, PhD, for her insightful contributions throughout its development.

## A PROMPTING

In this Section, we include the prompting strategies used in the experiments. The arguments `<case>, <question>,<options>, <explanation *>` are filled with content from the ESAP 2021-2022 program.

### A.1 Prompts in Experiment 1

**Prompt A**

‘Given the following clinical case, consider the question, and provide the most likely answer among the provided options and the reasoning process behind your choice.

~~~
<description>
<question>
Options: options
Please, reduce the output to a paragraph and report the logic steps that led to your answer.’
~~~

**Prompt B**

~~~
‘Consider this clinical case, question, and options available.
<description>
<question>
Options: <options>
Please do provide your choice among the possible answers. Then provide the reasoning steps behind your choice.’
~~~

### A.2 Prompt in Experiment 3

**Prompt**

~~~
‘Given the case the following clinical case and multiple choice question, I will provide you two potential
explanations.
Can you please report the level of agreement from a scale from 0 (null agreement) to 5 (perfect agreement) and
select which response makes more clinical sense between the two explanations?
Clinical case: <description>
Questions: <question Options: <options>
Explanation 1: <explanation 1>
Explanation 2: <explanation 2>
Please state clearly which is the most clinical correct option between explanation 1 and explanation 2. Then specify the agreement level between explanation 1 and explanation 2.’
~~~

In this case, `<explanation *>` will include one between the two reasoning paths: the gold-standard explanation of the ESAP program or the previous output of the LLM under consideration from Experiment 1.

## B EXPERIMENTAL SETUP

### B.1 Inference and Hyperparameters

We perform the model inference sequentially, with each model loaded once and used to process the full set of 91 clinical cases in a single session, each as a single batch. By calling the function for output generation separately for each prompt, we assure that the model output is independent from previous prompts. All the experiments are performed using AutoTokenizer+AutoModelForCausalLM+.generate() from the transformers library.

#### B.1.1 Experiment 1

do_sample set to False, temperature, top_p, and top_k set to None, length_penalty = 1.0, repetition_penalty = 1.2, max_tokens=1500, max_new_tokens = 1500, num_return_sequences = 1. pad_token_id = 128009, eos_token_id = 128009, and attention mask are output of the default tokenizer setting, with padding=True and truncation=True. All other arguments are left as default from the AutoTokenizer+AutoModelForCausalLM+.generate().

#### B.1.2 Experiment 2

Same hyperparameters of Experiment 1, with exception of do_sample = True, temperature=0.6, top_p=0.9, top_k=75.

#### B.1.3 Experiment 3

This shares identical hyperparameters of Experiment 1, with exception of max_new_tokens = 2048.

## C OUTPUT EVALUATION

## D VISUALIZATION OF MODEL CONSISTENCY ACROSS TEMPERATURES

Figure 3 shows how model correctness changes across different temperatures, for the three *T* values tested. ESAP items are ordered based on each model performance at the lowest temperature tested (*T* = 0.3). HuatuoGPT-o1-8B exhibits the highest coherence. Diabetica-7B and Diabetica-o1 represent the intermediate case. Interestingly, Diabetica-7B shows much higher consistency and correctness for *T* = 0.3 but its performance degrade quite abruptly for increasing temperature. Meditron3-8B, as discussed in the main body of the paper, avoid providing a response for *T* = 1.0, which explains the lack of higher numerical values.

## E FURTHER RESULTS FOR EXPERIMENT 3: DISCERNING GOLD-STANDARD REASONING

### E.1 Agreement between Gold Standard and Models Outputs from a Model’s Perspective

We extracted agreement scores from the model outputs on a 0–5 scale. For each case, we calculated the average agreement by combining the scores of two runs: one in which the gold standard explanation was presented first and the model’s reasoning second, and another with the order reversed. Since agreement is less informative when the model already selects the correct answer, our analysis focuses on cases where the model’s selected option disagreed with the gold standard choice. The goal of the following evaluation is to observe whether the level of agreement reflects the discrimination capabilities of the model to select the the gold standard explanation against the model’s own reasoning. We excluded the clinical cases where the model’s reasoning selects a different answer from the one in explanation 1 and explanation 2.

Figure 4 and Figure 5 show respectively the average agreement score for HuatuoGPT-o1-8B and Diabetica-o1.

The analysis can be only speculative given the small sample size. While Diabetica-o1 does not show any specific trend, the box plot distributions for HuatuoGPT-o1-8B suggests something interesting. In Figure 4, agreement scores are more widely distributed for cases where the model selected the gold standard explanation in both runs (Both), indicating greater variability of the agreement score and the potential capability of discerning clinical differences between the gold standard and the the misleading response of the model itself. Interestingly, when the model favored its own reasoning over the gold standard (Never) (*n* = 8 in prompt 1 and *n* = 4 in prompt 2), the box plot are generally more skewed towards higher agreement values, potentially reflecting a lack of sensitivity to the subtle distinctions in the gold standard explanation versus the model reasoning. The values in right plot (Never) of Figure 4a and Figure 4b can be qualitatively compared to the scores in Table 3, where the evaluation based on clinical reasoning is on a 1-5 scale. HuatuoGPT-o1-8B shows higher level of agreement than the ones assigned by the pediatric endocrinology expert. Evaluation of experiment 1 outcomes are given by the pediatric endocrinologist and HuatuoGPT-o1-8B (from Experiment 3).

### E.2 Integral Evaluations from Pediatric Endocrinologist Evaluation and HuatuoGPT-o1-8B

Table 9 and Table 10 show the clinician’s evaluation of HuatuoGPT-o1-8B responses of Experiment 1 and their evaluation in a Likert scale (1-5) based on the clinical reasoning scale in CLEVER. The right side of the tables show the evaluation of HuatuoGPT-o1-8B’s previous explanation against the gold standard. In general, we notice that HuatuoGPT-o1-8B tends to be more forgiving, losing some important details related to biochemical and genetic mechanisms which are key for the diagnosis.

**Table 4.**
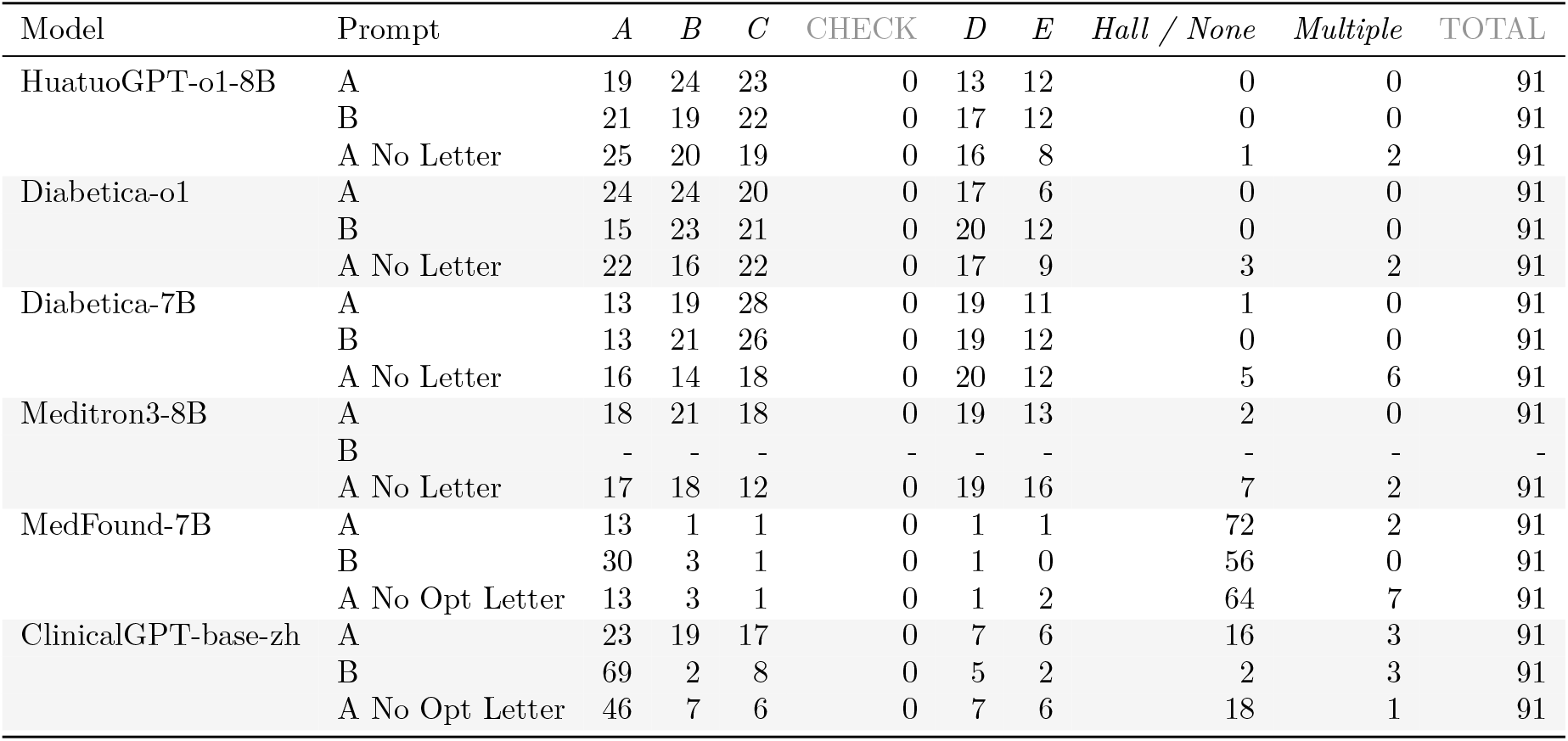
Experiment 1: Responses distribution for the three prompts tested. MedFound-7B and ClinicalGPT-base-zh show the highest degree of hallucination. Meditron3-8B does not output a response under prompt B.

**Table 5.** Experiment 2: Responses distribution across ten runs of prompt A.

| Model | Temperature | *A* | *B* | *C* | CHECK | *D* | *E* | *Hall / None* | *Multiple* | TOTAL |
|---|---|---|---|---|---|---|---|---|---|---|
| HuatuoGPT-o1-8B | 0.3 | 172 | 236 | 206 | 0 | 168 | 120 | 5 | 3 | 910 |
|  | 0.6 | 171 | 231 | 201 | 0 | 171 | 124 | 12 | 0 | 910 |
|  | 1.0 | 171 | 234 | 196 | 0 | 170 | 120 | 17 | 2 | 910 |
| Diabetica-o1 | 0.3 | 203 | 210 | 219 | 0 | 172 | 106 | 0 | 0 | 910 |
|  | 0.6 | 204 | 210 | 215 | 0 | 173 | 97 | 11 | 0 | 910 |
|  | 1.0 | 181 | 213 | 224 | 0 | 175 | 98 | 12 | 7 | 910 |
| Diabetica-7B | 0.3 | 146 | 170 | 276 | 0 | 194 | 113 | 8 | 3 | 910 |
|  | 0.6 | 170 | 166 | 252 | 0 | 198 | 110 | 12 | 2 | 910 |
|  | 1.0 | 158 | 169 | 237 | 0 | 162 | 123 | 53 | 8 | 910 |
| Meditron3-8B | 0.3 | 127 | 195 | 202 | 0 | 195 | 164 | 26 | 1 | 910 |
|  | 0.6 | 149 | 202 | 182 | 0 | 182 | 156 | 39 | 0 | 910 |
|  | 1.0 | 82 | 111 | 109 | 0 | 70 | 56 | 476 | 6 | 910 |

**Table 6.** Frequency of the majority class for *T* = 0.3.

|  | *A* | *B* | *C* | *D* | *E* | No Majority |
|---|---|---|---|---|---|---|
| HuatuoGPT-o1-8B | 17 | 24 | 17 | 16 | 10 | 7 |
| Diabetica-o1 | 18 | 20 | 22 | 13 | 8 | 10 |
| Diabetica-7B | 13 | 18 | 29 | 17 | 9 | 5 |
| Meditron3-8B | 11 | 19 | 20 | 17 | 13 | 11 |

**Table 7.** Frequency of majority class for *T* = 0.6.

|  | *A* | *B* | *C* | *D* | *E* | No Majority |
|---|---|---|---|---|---|---|
| HuatuoGPT-o1-8B | 16 | 22 | 21 | 17 | 12 | 3 |
| Diabetica-o1 | 22 | 18 | 23 | 16 | 7 | 5 |
| Diabetica-7B | 16 | 15 | 27 | 19 | 9 | 5 |
| Meditron3-8B | 9 | 23 | 16 | 17 | 11 | 15 |

**Table 8.** Frequency of majority class for *T* = 1.0. Meditron3-8B avoid option selection for most cases (71 out of 91).

|  | *A* | *B* | *C* | *D* | *E* | *Hall / None* | No Majority |
|---|---|---|---|---|---|---|---|
| HuatuoGPT-o1-8B | 17 | 23 | 20 | 15 | 11 | 0 | 5 |
| Diabetica-o1 | 13 | 19 | 25 | 16 | 8 | 0 | 10 |
| Diabetica-7B | 14 | 19 | 26 | 15 | 11 | 0 | 6 |
| Meditron3-8B | 3 | 4 | 0 | 2 | 2 | 71 | 9 |

**Table 9.**
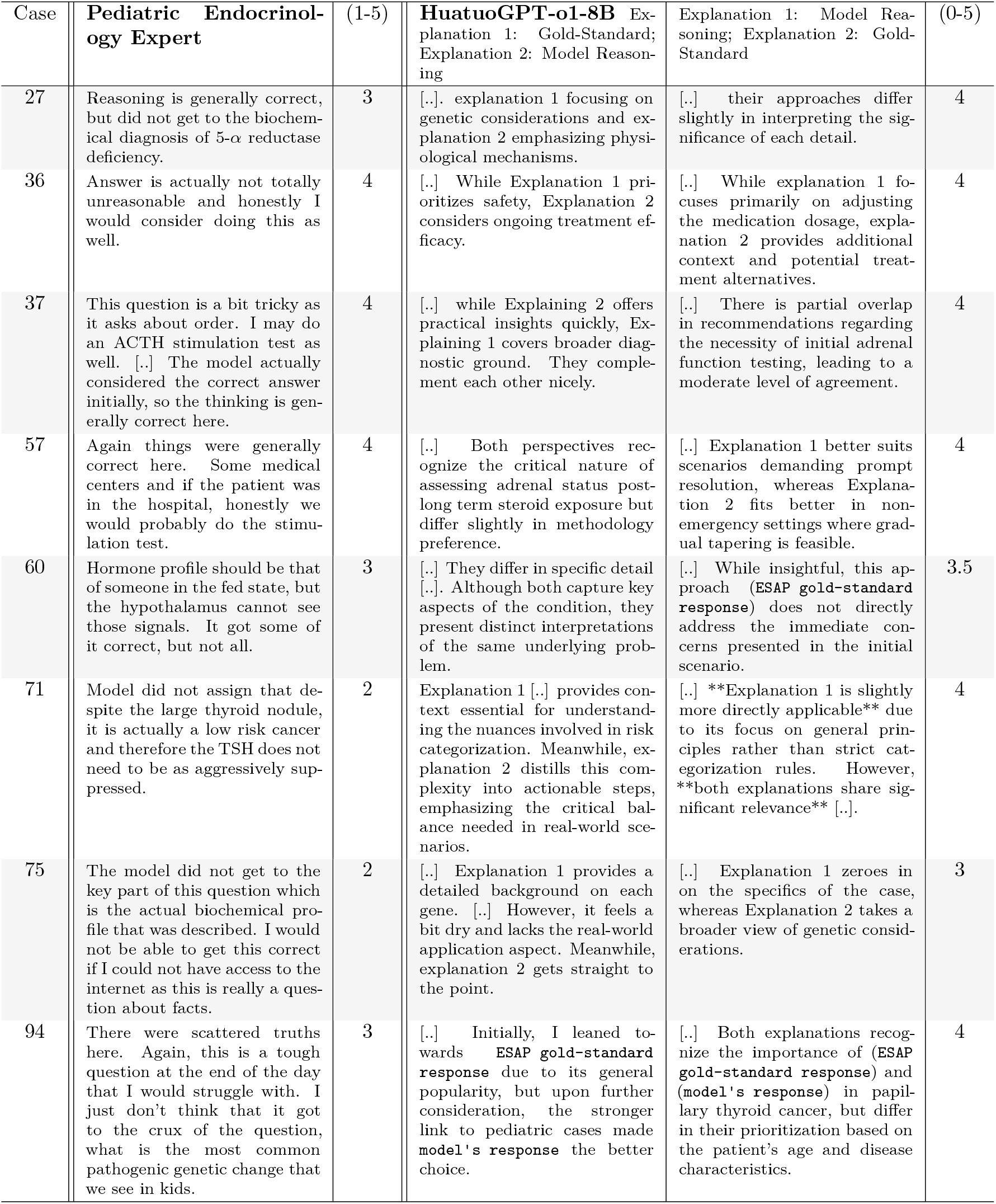
HuatuoGPT-o1-8B reasoning for incorrect answers to prompt 1 where model consistently failed to identify gold-standard reasoning (both). On the left, pediatric endocrinology expert evaluation, on the right, Huatuo-o1-8B evaluation.

| Case | **Pediatric Endocrinol-ogy Expert** | (1-5) | **HuatuoGPT-o1-8B** Ex-planation 1: Gold-Standard; Explanation 2: Model Reason-ing | Explanation 1: Model Rea-soning; Explanation 2: Gold-Standard | (0-5) |
|---|---|---|---|---|---|
| 27 | Reasoning is generally correct, but did not get to the biochem-ical diagnosis of 5-α reductase deficiency. | 3 | [..]. explanation 1 focusing on genetic considerations and ex-planation 2 emphasizing physi-ological mechanisms. | [..] their approaches differ slightly in interpreting the sig-nificance of each detail. | 4 |
| 36 | Answer is actually not totally unreasonable and honestly I would consider doing this as well. | 4 | [..] While Explanation 1 pri-oritizes safety, Explanation 2 considers ongoing treatment ef-ficacy. | [..] While explanation 1 fo-cuses primarily on adjusting the medication dosage, expla-nation 2 provides additional context and potential treat-ment alternatives. | 4 |
| 37 | This question is a bit tricky as it asks about order. I may do an ACTH stimulation test as well. [..] The model actually considered the correct answer initially, so the thinking is gen-erally correct here. | 4 | [..] while Explaining 2 offers practical insights quickly, Ex-plaining 1 covers broader diag-nostic ground. They comple-ment each other nicely. | [..] There is partial overlap in recommendations regarding the necessity of initial adrenal function testing, leading to a moderate level of agreement. | 4 |
| 57 | Again things were generally correct here. Some medical centers and if the patient was in the hospital, honestly we would probably do the stimu-lation test. | 4 | [..] Both perspectives rec-ognize the critical nature of assessing adrenal status post-long term steroid exposure but differ slightly in methodology preference. | [..] Explanation 1 better suits scenarios demanding prompt resolution, whereas Explana-tion 2 fits better in non-emergency settings where grad-ual tapering is feasible. | 4 |
| 60 | Hormone profile should be that of someone in the fed state, but the hypothalamus cannot see those signals. It got some of it correct, but not all. | 3 | [..] They differ in specific detail [..]. Although both capture key aspects of the condition, they present distinct interpretations of the same underlying prob-lem. | [..] While insightful, this ap-proach (ESAP gold-standard response) does not directly address the immediate con-cerns presented in the initial scenario. | 3.5 |
| 71 | Model did not assign that de-spite the large thyroid nodule, it is actually a low risk cancer and therefore the TSH does not need to be as aggressively sup-pressed. | 2 | Explanation 1 [..] provides con-text essential for understand-ing the nuances involved in risk categorization. Meanwhile, ex-planation 2 distills this com-plexity into actionable steps, emphasizing the critical bal-ance needed in real-world sce-narios. | [..] **Explanation 1 is slightly more directly applicable** due to its focus on general prin-ciples rather than strict cat-egorization rules. However, **both explanations share sig-nificant relevance** [..]. | 4 |
| 75 | The model did not get to the key part of this question which is the actual biochemical pro-file that was described. I would not be able to get this correct if I could not have access to the internet as this is really a ques-tion about facts. | 2 | [..] Explanation 1 provides a detailed background on each gene. [..] However, it feels a bit dry and lacks the real-world application aspect. Meanwhile, explanation 2 gets straight to the point. | [..] Explanation 1 zeroes in on the specifics of the case, whereas Explanation 2 takes a broader view of genetic consid-erations. | 3 |
| 94 | There were scattered truths here. Again, this is a tough question at the end of the day that I would struggle with. I just don’t think that it got to the crux of the question, what is the most common pathogenic genetic change that we see in kids. | 3 | [..] Initially, I leaned to-wards ESAP gold-standard response due to its general popularity, but upon further consideration, the stronger link to pediatric cases made model's response the better choice. | [..] Both explanations recog-nize the importance of (ESAP gold-standard response) and (model's response) in papil-lary thyroid cancer, but differ in their prioritization based on the patient’s age and disease characteristics. | 4 |

**Table 10.**
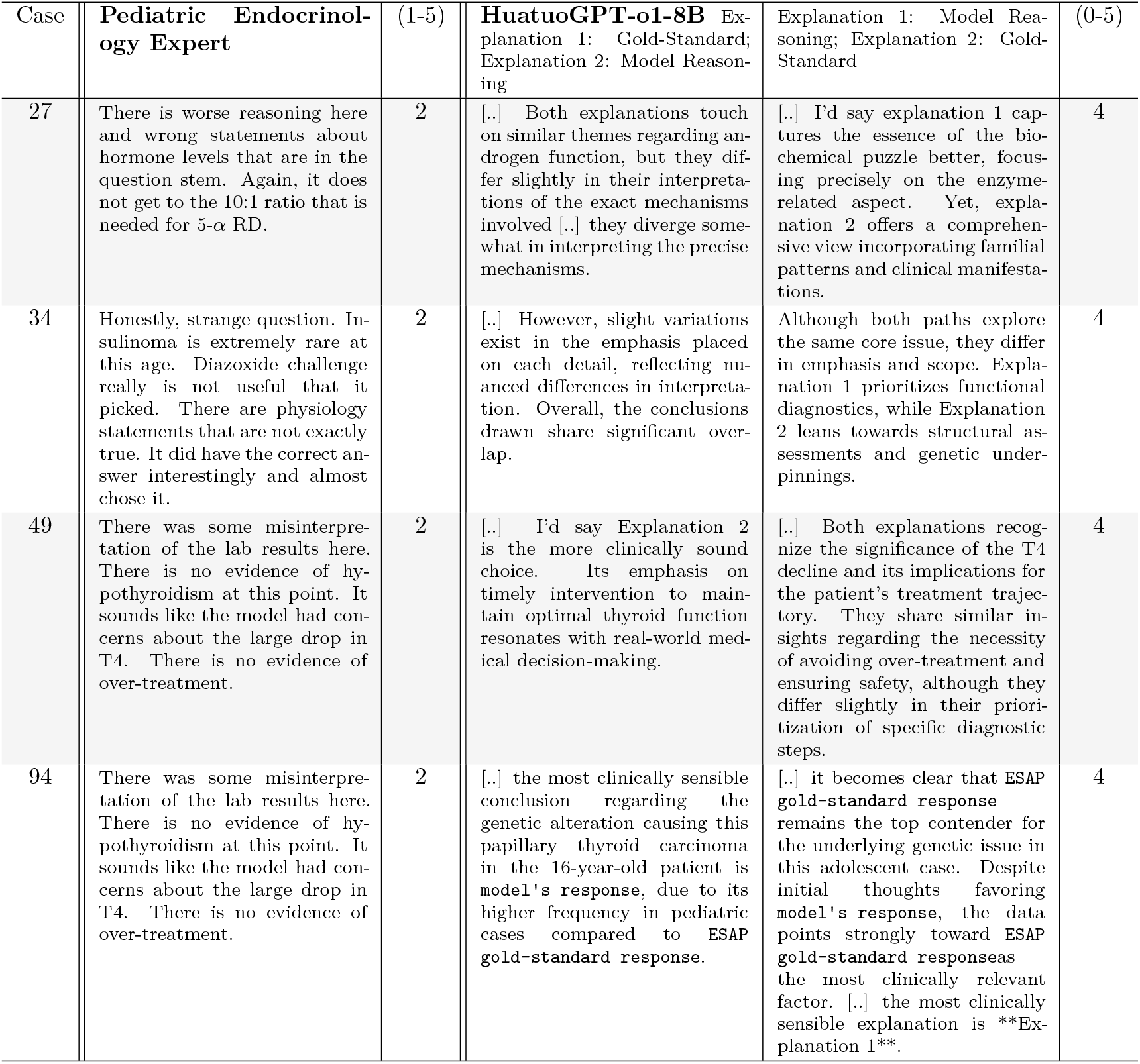
HuatuoGPT-o1-8B reasoning for incorrect answers to prompt 2 where model consistently failed to identify gold-standard reasoning (both). On the left, pediatric endocrinology expert evaluation, on the right, Huatuo-o1-8B evaluation.

| Case | **Pediatric Endocrinology Expert** | (1-5) | **HuatuoGPT-o1-8B** Explanation 1: Gold-Standard; Explanation 2: Model Reasoning | Explanation 1: Model Reasoning; Explanation 2: Gold-Standard | (0-5) |
|---|---|---|---|---|---|
| 27 | There is worse reasoning here and wrong statements about hormone levels that are in the question stem. Again, it does not get to the 10:1 ratio that is needed for 5-α RD. | 2 | [..] Both explanations touch on similar themes regarding androgen function, but they differ slightly in their interpretations of the exact mechanisms involved [..] they diverge somewhat in interpreting the precise mechanisms. | [..] I’d say explanation 1 captures the essence of the biochemical puzzle better, focusing precisely on the enzyme-related aspect. Yet, explanation 2 offers a comprehensive view incorporating familial patterns and clinical manifestations. | 4 |
| 34 | Honestly, strange question. Insulinoma is extremely rare at this age. Diazoxide challenge really is not useful that it picked. There are physiology statements that are not exactly true. It did have the correct answer interestingly and almost chose it. | 2 | [..] However, slight variations exist in the emphasis placed on each detail, reflecting nuanced differences in interpretation. Overall, the conclusions drawn share significant overlap. | Although both paths explore the same core issue, they differ in emphasis and scope. Explanation 1 prioritizes functional diagnostics, while Explanation 2 leans towards structural assessments and genetic underpinnings. | 4 |
| 49 | There was some misinterpretation of the lab results here. There is no evidence of hypothyroidism at this point. It sounds like the model had concerns about the large drop in T4. There is no evidence of over-treatment. | 2 | [..] I’d say Explanation 2 is the more clinically sound choice. Its emphasis on timely intervention to maintain optimal thyroid function resonates with real-world medical decision-making. | [..] Both explanations recognize the significance of the T4 decline and its implications for the patient’s treatment trajectory. They share similar insights regarding the necessity of avoiding over-treatment and ensuring safety, although they differ slightly in their prioritization of specific diagnostic steps. | 4 |
| 94 | There was some misinterpretation of the lab results here. There is no evidence of hypothyroidism at this point. It sounds like the model had concerns about the large drop in T4. There is no evidence of over-treatment. | 2 | [..] the most clinically sensible conclusion regarding the genetic alteration causing this papillary thyroid carcinoma in the 16-year-old patient is model's response, due to its higher frequency in pediatric cases compared to ESAP gold-standard response. | [..] it becomes clear that ESAP gold-standard response remains the top contender for the underlying genetic issue in this adolescent case. Despite initial thoughts favoring model's response, the data points strongly toward ESAP gold-standard responseas the most clinically relevant factor. [..] the most clinically sensible explanation is **Explanation 1**. | 4 |

## F PIPELINE VS AUTOTOKENIZER + AUTOMODELFORCAUSALLM.GENERATE()

In this study, all small open-source LLMs were run using the HuggingFace `transformers` library. Text generation was performed using two approaches: the high-level `pipeline()` interface and a lower-level setup involving `AutoTokenizer, AutoModelForCausalLM`, and manual use of `.generate()`. Both were configured with `do_sample=False` and a `temperature=0`, which should theoretically result in deterministic result. Between the two workflows, only the pad_token_id and eos_token_id parameters are different, but our understanding is that, since we avoid the use of batches and call the function for output generation separately for each prompt, those tokens should not affect the model generation.

Although in some cases the two methods produced identical responses, we observed instances where the generated outputs diverged after the initial few tokens, despite identical prompts and generation settings. In the manual `.generate()` setup, we explicitly set `pad_token_id` and `eos_token_id` to the model’s end-of-sequence token.

However, these parameters are not exposed or controlled in the same way through `pipeline()`, which can lead to implicit differences in how padding or attention masks are handled during inference.

This divergence highlights how small numerical variations might cause significant differences in auto-regressive generation, even under deterministic decoding conditions. This invites further considerations when evaluating model consistency and results reproducibility.

**AutoTokenizer + AutoModelForCausalLM**

\## Thinking

Alright, let’s think about this little guy who just came in with some pretty interesting symptoms. So first off, we’ve got him having trouble with his eyes - specifically, there’s something going on with his vision where one eye turns inward. That happened right after he took quite a tumble when he fell while trying to stand up by himself. Now, they did an MRI and found that both of his optic nerves look swollen, kind of like they’re twisted around too much, and even the back part of his eyeballs seem a bit flat. This all sounds like it could be related to increased pressure inside his skull, maybe because of fluid buildup or swelling somewhere.

Now, moving onto what else is happening with him… Oh, I see! *They mentioned he has an unusual head shape. It seems like his skull isn’t developing as expected, especially across the top where you’d expect two separate parts but instead, everything fused together into more of a long oval shape. And then there’s how tall he is – way shorter than other kids his age, definitely below the fifth percentile. Plus, he’s got these issues with his legs, like bowing and a funny walk. Hmm, those sound familiar*…

[..]

\## Final Response

*The clinical findings described in the scenario point towards a skeletal disorder characterized by disproportionate short stature, limb deformities, and craniofacial anomalies. Given the presence of short stature (< 5*^*th*^ *percentile), mild genu varum (bow-leggedness) deformity, and a waddling gait, along with the distinct head shape indicating complete fusion of the sagittal suture and dolichocephaly*, ***the most likely underlying skeletal disorder is **C. Hypochondroplasia\*\****. *This diagnosis aligns with the characteristic features observed in the patient, particularly the absence of dental abnormalities and the nature of the limb deformities. Other potential diagnoses, such as vitamin D deficiency or genetic syndromes, do not fully account for the specific constellation of symptoms presented. Therefore, hypochondroplasia emerges as the most plausible explanation for the child’s condition*.

**Pipeline**

\## Thinking

Alright, let’s think about this little guy who just came in with some pretty interesting symptoms. So first off, we’ve got him having trouble with his eyes - specifically, there’s something going on with his vision where one eye turns inward. That happened right after he took quite a tumble when he fell while trying to stand up by himself. Now, they did an MRI and found that both of his optic nerves look swollen, kind of like they’re twisted around too much, and even the back part of his eyeballs seem a bit flat. This all sounds like it could be related to increased pressure inside his skull, maybe because of fluid buildup or swelling somewhere.

Now, moving onto what else is happening with him… Oh, I see! *There’s more than just the eyes. They mentioned he has a funny-shaped head – not round but sort of long and narrow, almost like a football. And then there’s how tall he isn’t growing compared to other kids his age. Plus, he’s got these bowed legs and walks with a weird, wobbly gait. It seems like everything points towards some bone issues here. Let me think*…

[..]

\## Final Response

*The clinical presentation described suggests several key features: bilateral eye issues post-fall, unusual cranial shape, short stature*,

*leg bowing, and a distinctive gait. Given these details*, ***the condition that aligns most closely with these findings is **D. Severe vitamin D deficiency\*\****. *This conclusion is supported by the presence of rachitic changes seen on x-ray, indicating poor bone mineralization typically caused by insufficient vitamin D intake during critical growth periods. While other conditions such as hypochondroplasia may share similar skeletal manifestations, the specific context of inadequate vitamin D supplementation early in life strongly supports severe vitamin D deficiency as the primary concern. Other options listed do not fully account for the comprehensive symptom set presented. Therefore, option D is the most fitting diagnosis based on the information provided*.

